# The effect of influenza vaccination on trained immunity: impact on COVID-19

**DOI:** 10.1101/2020.10.14.20212498

**Authors:** Priya A. Debisarun, Patrick Struycken, Jorge Domínguez-Andrés, Simone J.C.F.M. Moorlag, Esther Taks, Katharina L. Gössling, Philipp N. Ostermann, Lisa Müller, Heiner Schaal, Jaap ten Oever, Reinout van Crevel, Mihai G. Netea

## Abstract

Every year, influenza causes 290.000 to 650.000 deaths worldwide and vaccination is encouraged to prevent infection in high-risk individuals. Interestingly, cross-protective effects of vaccination against heterologous infections have been reported, and long-term boosting of innate immunity (also termed *trained immunity*) has been proposed as the underlying mechanism. Several epidemiological studies also suggested cross-protection between influenza vaccination and COVID-19 during the current pandemic. However, the mechanism behind such an effect is unknown. Using an established *in-vitro* model of trained immunity, we demonstrate that the quadrivalent inactivated influenza vaccine used in the Netherlands in the 2019-2020 influenza season can induce a trained immunity response, including an improvement of cytokine responses after stimulation of human immune cells with SARS-CoV-2. In addition, we found that SARS-CoV-2 infection was less common among Dutch hospital employees who had received influenza vaccination during the 2019/2020 winter season (RR = 0,61 (95% CI, 0.4585 - 0.8195, *P* = 0.001). In conclusion, a quadrivalent inactivated influenza vaccine can induce trained immunity responses against SARS-CoV-2, which may result in relative protection against COVID-19. These data, coupled with similar recent independent reports, argue for a beneficial effect of influenza vaccination against influenza as well as COVID-19, and suggests its effective deployment in the 2020-2021 influenza season to protect against both infections.

## INTRODUCTION

As of October 2020 there were over 37 million confirmed cases and one million deaths due to COVID-19 [1]. In many cases, SARS-CoV-2 infections only cause mild symptoms that resolve spontaneously. However, in the elderly or in patients with underlying health conditions such as cardiovascular disease, obesity, diabetes or pre-existing lung conditions, the disease is often more severe and potentially lethal. Various complications can arise and include, pulmonary edema, severe pneumonia, acute respiratory distress syndrome (ARDS) and thrombotic complications among others [2]. Due to the rapid spread and the high clinical and socio-economic burden of COVID-19, the efforts to prevent and combat the disease have been enormous. Despite the numerous ongoing developments and clinical trials to create specific vaccines against the virus, the earliest expected vaccine is likely to be deployed at least 4-6 months from now and is not expected to be readily available on a large scale [3, 4].

In addition to COVID-19, we are still exposed to other threatening pathogens. In countries with temperate climates, seasonal influenza outbreaks mainly occur in the winter, with the first cases starting to appear as early as September. This leads to recurrent widespread mortality and morbidity causing 3 to 5 million cases of severe illness and 290.000 to 650.000 deaths per year [5]. During the previous 2017/2018 flu season in Europe, an estimated excess mortality of 125.000 deaths was measured [6]. Because of the high morbidity and socioeconomic burden of recurrent influenza epidemics, vaccination has become a key strategy in protecting high-risk individuals against the flu and is therefore a widely promoted public health strategy [7-10]. Despite the wide use of flu vaccines, there is contradictory information on how the influenza vaccine might affect the outcome of other infections, including COVID-19. The potential interaction between vaccines and infections other than their target disease, has attracted a great deal of attention lately. It has been demonstrated that certain vaccines (such as bacillus Calmette-Guérin (BCG), measles-containing vaccines, or oral polio vaccine) have strong beneficial protective effects through long-term boosting of innate immunity, a process called *trained immunity* [11]. In line with this, several recent studies suggested a potential beneficial effect of influenza vaccination on susceptibility to COVID-19 [12, 13]. Despite earlier reports that have shown little or opposite effects of influenza vaccines on heterologous infections in children [14-19]. With the flu season on its way and influenza vaccination campaigns starting off soon, it is paramount to clarify the exact effects of influenza vaccination on the incidence and the disease course of COVID-19.

In this study, we investigated the possible induction of trained immunity responses by the influenza vaccine used in the 2019-2020 winter season in the Netherlands. In addition, we assessed the correlation between influenza vaccination, the incidence of COVID-19, and the disease outcome, using the influenza vaccination rates in employees of the Radboud University Medical Center, one of the large academic hospitals of the Netherlands.

## METHODS

### Blood donors

Buffy coats from healthy adult donors were obtained after written informed consent (Sanquin blood bank, Nijmegen, The Netherlands). The study was approved by the Arnhem-Nijmegen Medical Ethical Committee.

### *In-vitro* influenza training model

Peripheral blood mononuclear cells (PBMCs) were isolated using Ficoll-Paque (VWR, Tingalpa, Australia) density gradient isolation and resuspended in RPMI 1640+ (Dutch modified, ThermoFisher, Waltham, MA, USA) culture medium supplemented with 50 mg/mL gentamicin, 2 mM glutamax (ThermoFisher), and 1 mM pyruvate (ThermoFisher). PBMCs were diluted to a concentration of 5*10^6^ cells/ml and 100μl (500.000 cells) of cell suspension was added to each well of a 96-wells round bottom plate. PBMCs were stimulated with Vaxigrip Tetra® (Sanofi Pasteur Europe), which is similar to the vaccine used in the 2019/2020 vaccination campaign in the Netherlands. The influenza vaccine contains 15 grams of Hemagglutinin each from 4 inactivated, non-adjuvanted virus strains (1 A/Guangdong-Maonan/SWL1536/2019 (H1N1)pdm09-like strain, (A/GuangdongMaonan/SWL1536/2019, CNIC-1909, 2 A/Hong Kong/2671/2019 (H3N2) –like strain, (A/Hong Kong/2671/2019, IVR-208), 3 B/Washington/02/2019 – like strain (B/Washington/02/2019, wild type), 4 B/Phuket/3073/2013-like strain (B/Phuket/3073/2013, wild type). Vaxigrip Tetra® was added to the wells in 10, 50, 100 and 400-fold dilutions in 1640 culture medium (RPMI medium; Invitrogen, CA, USA) supplemented with 10% human pooled serum. In addition, 5 μg/ml of BCG (Bacille Calmette-Guérin, SSI, Denmark) was added to half of the wells to investigate the potency of influenza vaccine to enhance trained immunity induced by BCG. PBMCs were incubated for 24h after which the supernatants were harvested. Cells were washed with warm PBS and medium was refreshed. PBMCs were then left to rest in culture medium for another 5 days, after which (on day 6) they were restimulated with LPS (10 ng/mL) or inactivated SARS-CoV-2 [20] (SARS-CoV-2 NRW-42 isolate, 40x diluted, TCID50/mL 6.67*10^4^) for 24 hours. On day 7, supernatants were harvested and stored at −20°C for cytokine measurements (Figure 1).

**Figure 1.**
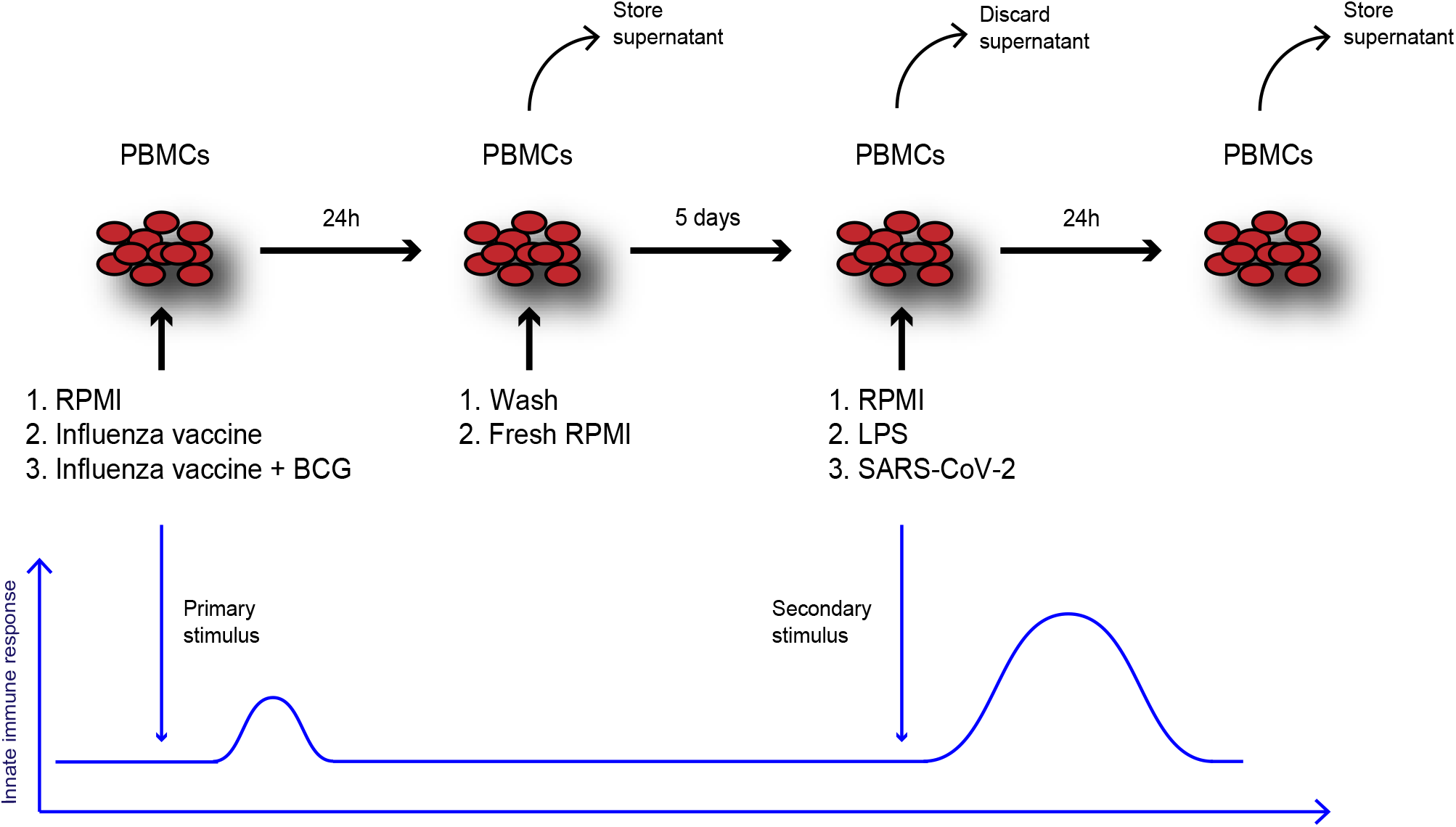
*In-vitro* trained immunity experiments with quadrivalent inactivated influenza vaccine and BCG. PBMCs from healthy donors were isolated and seeded in a 96-wells round bottom plate. Then 4 dilutions (10x, 50x, 100x, 400x) of the quadrivalent inactivated influenza vaccine (Vaxigrip Tetra®) were added to the cells. In other wells, the vaccine was combined with BCG (5µg/mL) and control wells only contained RPMI. After 24 h the supernatant was collected and cells were washed. Fresh RPMI was added to the cells and left to incubate for another 5 days. On day 6, the PBMCs were restimulated with LPS (10 ng/mL) or inactivated SARS-CoV-2 (40x diluted, TCID50/mL 6.67*10e4) for another 24 h. On day 7, the supernatants were collected and stored for cytokine measurements.

### Lactate-dehydrogenase (LDH) assay

Cytotoxicity was measured using LDH concentrations in fresh supernatants collected after 24 h stimulation, using the Cyto-Tox96 Non-Radioactive cytotoxicity assay (Promega, WI, USA).

### Cytokine measurements

Cytokine concentrations were determined in 24 h supernatants (TNF-*α*, IL-6, IL-1β) and 7 day supernatants (TNF-*α*, IL-6, IL-1RA, and IFN-γ) using commercial ELISA kits (R&D systems, Bio-Techne, Minneapolis, Minnesota, USA) according to the manufacturer’s protocol.

### Observational data healthcare workers

The Radboudumc hospital registration database of SARS-CoV-2 PCR-positive healthcare workers as of June 1^st^ 2020 was consulted. The corresponding influenza vaccination status of healthcare workers was retrieved from the database of the Department of Occupational Health and Safety of the hospital, as well as the total influenza vaccination coverage rate (VCR) of the Radboudumc during the flu season of 2019/2020. Additionally, SARS-CoV-2 positive employees were sent a questionnaire to assess disease duration, severity and comorbidities. Disease duration was measured as the number of days between the SARS-CoV-2 PCR test and the first day employees resumed their work. All hospital employees are equally offered an influenza vaccination every year. However, SARS-CoV-2 testing in the beginning of the pandemic was only available for employees who were indispensable for patientcare, due to shortage of testing materials. Giving the observational nature of this study and the use of short questionnaires only, no ethical approval was required.

### Data analysis

Hospital database analysis was done using IBM SPSS statistics 25. To assess the association between COVID-19 incidence and influenza vaccination status, a Chi-square test was used. No correction for confounding was possible because no individual characteristics were available in the SARS-CoV-2 PCR negative health care workers; only influenza vaccination status data for the entire group was known. Missing values of other variables were left out of analysis. Cytokine and LDH concentrations were analyzed using Wilcoxon matched-pairs signed rank test. Absolute cytokine concentrations were determined after 24 hours in influenza vaccine stimulated conditions (with and without BCG) and compared with unstimulated conditions. As a readout for trained immunity responses, influenza vaccine-primed (at day 6 LPS and SARS-CoV-2 restimulated) conditions were compared to RPMI conditions as a negative control and BCG only conditions as a positive control. To assess synergistic effects on trained immunity, combined influenza vaccine and BCG stimulated conditions were also compared. LDH values were calculated to percentages of cell death, with lysed cells as a positive control. Data were analyzed using Graphpad 8.02 (La Jolla, San Diego, CA, USA). A two-sided P-value below .05 (*) or below .01 (**) was considered statistically significant. Data are shown as means ± SEM.

## RESULTS

### A quadrivalent inactivated influenza vaccine (Vaxigrip Tetra®) amplifies cytokine production in human PBMCs

Freshly isolated PBMCs from healthy donors (n=9) were stimulated for 24 h with Vaxigrip Tetra® in 10, 50, 100, or 400 times dilutions alone or in presence of BCG. Stimulation with Vaxigrip Tetra® alone did not result in increased production of IL-6, TNF-*α*, or IL-1β by itself. However, the combination of the influenza vaccine with BCG induced significantly higher cytokine production compared to BCG alone, suggesting a synergistic effect between these two vaccines. This effect was dose-dependent, with higher dilutions of the influenza vaccine associated with a higher cytokine production (Figure 2). To rule out cell death as a cause of cytokine release, LDH concentrations were measured in fresh supernatants collected after 24 h stimulation and showed no differences between conditions (Supplementary Figure 1), arguing against toxic effects of the vaccines.

**Figure 2.**
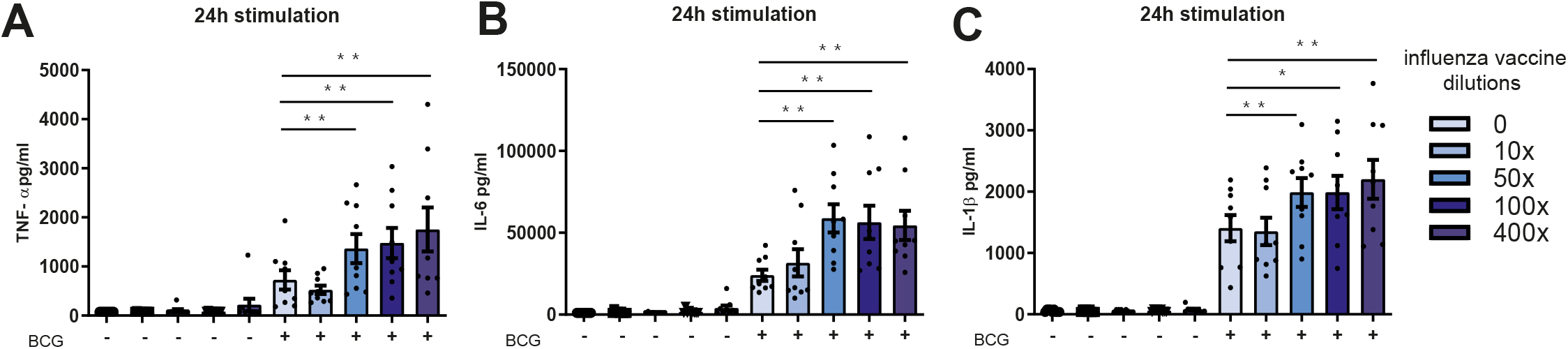
Addition of quadrivalent inactivated influenza vaccine increases 24 h cytokine production in PBMCs stimulated with BCG. Stimulation of PBMCs with several dilutions of the quadrivalent influenza vaccine (10x, 50x, 100x, 400x) did not result in increased concentrations of IL-6, IL-1β, and TNF-α after 24 h compared to RPMI conditions. Vaxigrip Tetra® increased cytokine production induced by BCG (5ug/ml). (Wilcoxon matched pairs signed rank test, *n* = 9, * = *P <* .05, ** = *P* <.01).

### Vaxigrip Tetra® induces trained immunity and amplifies BCG-induced trained immunity

In a separate set of experiments, 6 days after an initial 24h period of training of human PBMCs with the quadrivalent inactivated influenza vaccine, the cells were restimulated with heat-inactivated SARS-CoV-2 or lipopolysaccharide (LPS) from *Escherichia coli* for another 24h. Training with different concentrations of the influenza vaccine induced the production of higher levels of IL-1RA after restimulation with SARS-CoV-2 for all the dilutions tested, as well as for LPS (400x dilution) (figure 3A, B). Combining the influenza vaccine with BCG abrogated this effect. Trained immunity responses on the production of IL-6 were observed in almost all influenza vaccine-primed conditions after restimulation with LPS (all dilutions) and heat-inactivated SARS-CoV-2 (10x, 100x, 400x dilutions) (Figure 3C). In addition, induction of trained immunity by BCG was amplified by training with the combination of BCG and Vaxigrip Tetra® (Figure 3D). PBMC training with the influenza vaccine alone also enhanced IFN-γ production stimulated with SARS-CoV-2, but not LPS (Figure 3E). The combination of BCG with Vaxigrip Tetra® also induced higher responses upon restimulation with SARS-CoV-2, but not with LPS (Figure 3F). None of the influenza vaccine-primed conditions produced any significant release of TNF-α after SARS-CoV-2 restimulation. Only in BCG-primed conditions after LPS restimulation a higher induction of TNF-α was present (Figure 3G, H). Overall, training with various concentrations of the influenza vaccine enhanced cytokine production upon restimulation, and also amplified the trained immunity inducing capacity of BCG.

**Figure 3.**
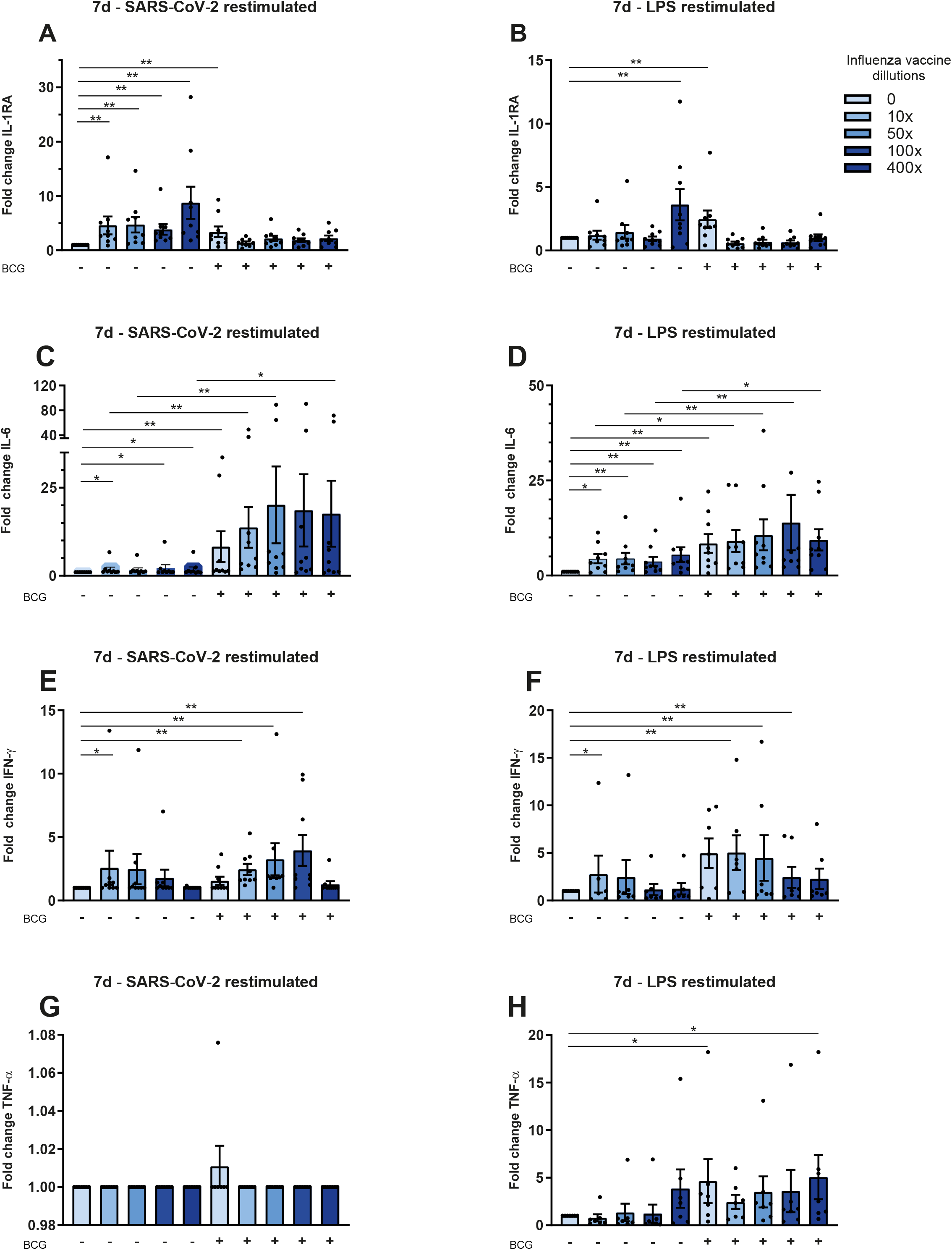
Inactivated quadrivalent influenza vaccine induces trained immunity after restimulation with SARS-CoV-2 and LPS and influences BCG-induced training. PBMCs were trained with several dilutions of the inactivated quadrivalent vaccine (10x, 50x, 100x, 400x) with or without BCG (5 μg/mL), restimulated at day 6 with LPS (10 ng/ml) or SARS-CoV-2 ((40x diluted, TCID50/mL 6.67*10e4) for 24 h and compared to RPMI conditions. Vaxigrip Tetra® amplified IL-1Ra (A, B) and IL-6 (C, D) responses after SARS-CoV-2 or LPS restimulation, compared to stimulation of naïve PBMCs. Training with BCG diminished the training effect of Vaxigrip Tetra® on IL-1Ra production, but increased IL-6 responses even more. A significant increase in IFN-γ production was seen after training of PBMCs with 10x dilutions of the influenza vaccine and restimulation with SARS-CoV-2. The combination of the influenza vaccine with BCG induced even more IFN-γ than BCG alone (E, F). No significant increase in TNF-α by Vaxigrip Tetra® was observed after restimulation with SARS-CoV-2 (G). Vaxigrip Tetra® enhanced TNF production upon LPS restimulation, but only in combination with BCG (H). (Wilcoxon matched pairs signed rank test, *n* =7-9, * = *P <* .05, ** = *P* <.01)

### Quadrivalent inactivated influenza vaccination is associated with lower COVID-19 incidence

As of June 1^st^ 2020, at the end of the first wave of the COVID-19 pandemic in the Netherlands, Radboud University Medical Center counted a total of 10.631 employees of which 184 were documented as SARS-CoV-2 PCR-positive according to the hospital’s database. The average age of the employees was 41 years and 42 years in the SARS-CoV-2 positive and negative group, respectively. Female employees made up a slightly higher proportion of the SARS-CoV-2 positive group (79%) compared to the SARS-CoV-2 negative group (70%) (Table 1).

**Table 1.** Baseline characteristics of SARS-CoV-2 negative and positive employees.

| Characteristic* | SARS-CoV-2 negative (n=10447) | SARS-CoV-2 positive (n=184) |
| --- | --- | --- |
| Gender, female† | 7338 (70,2) | 146 (79,3) |
| Gender, male † | 2986 (28,6) | 39 (21,2) |
| Age, years † | 42 ± 12,9 | 41 ± 12,3 |
| Direct patient-contact | 4412 (42,2) | 139 (75,5) |

\* Mean ± SD for continuous variables, and N (%) for categorical variables
† Age and gender were not provided for 123 employees in the SARS-CoV-2 negative group

To investigate the effect of influenza vaccination on the incidence of COVID-19, we gathered the influenza vaccination status of SARS-CoV-2 PCR positive employees in the Radboud University Medical Center and compared it to the influenza vaccination coverage rates in hospital employees unaffected by COVID-19. Within the SARS-CoV-2 positive group, 42% (77/184) of the individuals were vaccinated with influenza during the flu season of 2019/2020, whereas the total influenza vaccine coverage rate for that season in SARS-CoV-2 negative personnel was 54% (5664/10447). This resulted in a 2.23% incidence of COVID-19 in the non-vaccinated individuals, while the incidence of COVID-19 in influenza vaccinated individuals was 1.33%: thus, a statistically significant negative association between influenza vaccination and COVID-19 incidence of RR = 0,61 (95% CI, 0.46 - 0.82, *P* = 0.001) (Χ^2^(1, N = 10632) = 11,41, p = .0008) (Figure 4A, B) was identified. We found no association between influenza vaccination status and COVID-19 duration: the mean disease duration in influenza-unvaccinated personnel was 17±9 days, and 18±11 days in vaccinated individuals (*P* = .23). It is important to note that 76% of SARS-CoV-2 positive employees had direct patient contact, as opposed to 42% in SARS-CoV-2 negative personnel.

**Figure 4.**
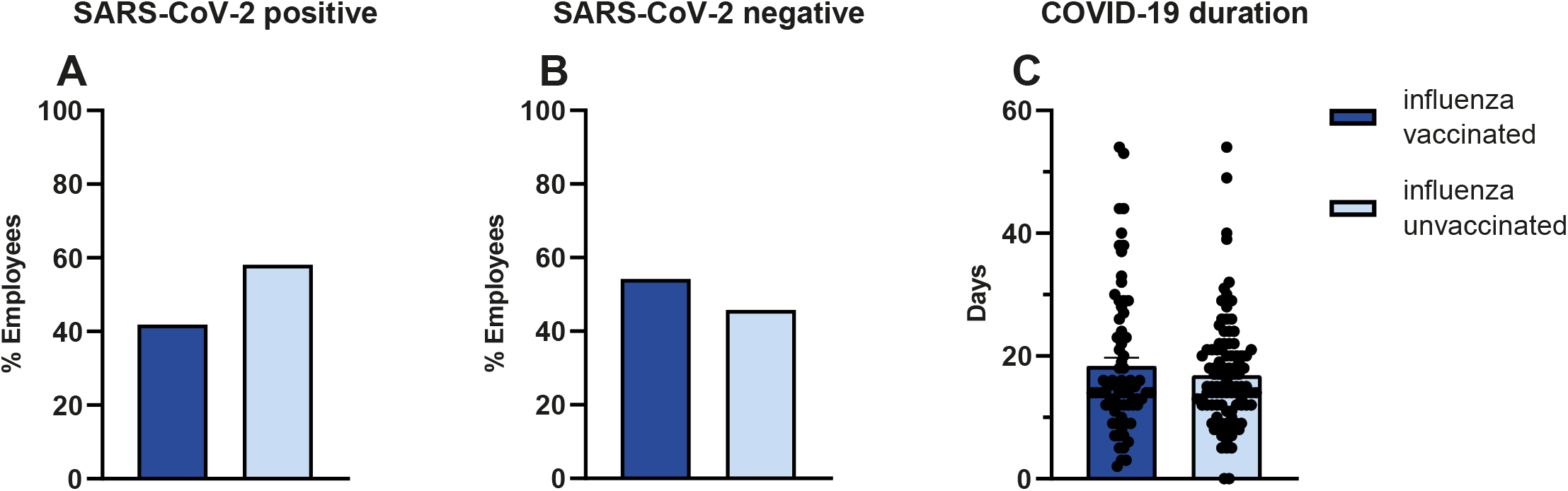
Influenza vaccination is associated with lower COVID-19 incidence. In SARS-CoV-2 positive employees, 42% (77/184) was influenza vaccinated. (A) In SARS-CoV-2 negative personnel 54% (5664/10447) was vaccinated, (B) Vaccination was associated with lower COVID-19 incidence RR = 0,61 (95% CI, 0.4585 - 0.8195, *P* = 0.001), (Χ^2^(1, N = 10632) = 11,41, *P =* .0008). No association was found between vaccination status and COVID-19 duration (Χ^2^(42, N = 172) = 48,41 p =.23). (C) The mean disease duration in influenza unvaccinated personnel was 17±9 days and 18±11 days in vaccinated individuals.

Among SARS-CoV-2 positive employees, only one of the individuals was hospitalized, but did not need any intensive-care treatment. No SARS-CoV-2 related deaths among positive employees occurred. The mean ages were 39 and 44 years for influenza unvaccinated and vaccinated employees within the SARS-CoV-2 positive group respectively. There were no significant differences between sex, patient contact or comorbidities in both groups (Table 2).

**Table 2.** Baseline characteristics of influenza vaccinated and unvaccinated employees.

| Characteristic* | SARS-CoV-2 positive |  |
| --- | --- | --- |
|  | Influenza unvaccinated (n=107) | Influenza vaccinated (n=77) |
| Gender, female | 85 (79,4) | 61 (79,2) |
| Gender, male | 22 (20,6) | 16 (20,1) |
| Age, years | 39 ± 12,2 | 44 ± 12,0 |
| Direct patient-contact | 77 (73,0) | 62 (80,6) |
| Total comorbidities † | 9 (20) | 10 (19) |

\* Mean ± SD for continuous variables, and N (%) for categorical variables
† comorbidities were not known for 58 influenza unvaccinated and 25 influenza vaccinated individuals

## DISCUSSION

In the present study, we demonstrate that the tetravalent inactivated influenza vaccine Vaxigrip Tetra® induces trained immunity in an established *in-vitro* model, resulting in improved responsiveness of immune cells to SARS-CoV-2 stimulation. In addition, Vaxigrip Tetra® amplifies the capacity of the BCG vaccine to induce trained immunity [21]. These results are in line with a previous study from our group in which 40 healthy volunteers received a trivalent influenza vaccine 14 days after receiving either BCG or placebo [22]. Whilst BCG exhibited a broad increase of pro-inflammatory cytokines for a larger set of unrelated pathogens, the influenza vaccine showed more selective augmentation of cytokine responses after *ex-vivo* restimulation of peripheral blood leukocytes, similar to our observations. In addition, we complement these data with an epidemiological analysis that shows an inverse association between influenza vaccination using a quadrivalent inactivated influenza vaccine, and COVID-19 incidence. This suggests a protective effect of the flu vaccine against infection with SARS-CoV-2.

Our data are in line with several recent ecological studies (Table 3). Hernandez et al. employed publicly available data from the 2019/2020 influenza vaccination season in Italy with a quadrivalent vaccine, a trivalent inactivated vaccine, and an inactivated, adjuvanted trivalent vaccine to calculate a linear regression model to predict the COVID-19 mortality in vaccinated adults over 65 years of age. They found a moderate to strong negative correlation (r = -.5874, n = 21, *P* = .0051) between influenza vaccination and mortality in the elderly, suggesting that if the influenza VCR was higher, less individuals died from COVID-19 [23]. Another Italian study used data from 21 Italian regions and also found a negative correlation between influenza VCR and SARS-CoV-2 seroprevalence, hospitalization rates, ICU admissions as well as COVID-19 related mortality in the elderly. Multivariable analysis revealed a *R*^2^ of 0.88, 0.82, 0.70 and 0.78 for all the outcomes respectively. In other words, influenza vaccination strongly predicts the variance in the aforementioned outcomes [13]. Zanettini et al. concluded that a 10% increase in VCR could decrease SARS-CoV-2 related mortality by 28 percent, adjusted for several variables [24]. Arokiaraj also described the existence of negative correlations between VCR and COVID-19 related morbidity and mortality in individuals over 65 years of age in members of the Organisation for Economic Cooperation and Development (OECD) countries. However, these correlations were not substantiated by statistical evidence, making it difficult to draw firm conclusions [25]. On the contrary, Lisewski et al. found an increased COVID-19 risk in 28 OECD countries as a result of influenza vaccination, assessed with a Pearson correlation coefficient of r = 0.58 (95CI: 0.27 to 0.78; p=0.001), between VCR and attack rates [26]. A recent report from the Evidence-based medicine, public health and environmental toxicology consortium (EBMPHET), compared the vaccination coverage rate among elderly (≥ 65 years of age) and COVID-19 infection risk and disease severity in Europe and the USA. They found a statistically significant positive correlation between the VCR and reported COVID-19 incidence in Europe (r = 0.66 ± 0.13, *P* = .000017) as well as mortality for Europe (r = 0.68 ± 0.13, p = 0.000006) and the USA, but confounding factors were not taken into account [27].

**Table 3.** Studies on the association between influenza vaccination and COVID-19 related outcomes.

| Author, year | Design | Methods | Main results |
| --- | --- | --- | --- |
| Hernandez et al., 2020 | Ecological study | COVID-19 mortality in elderly (> 65 years) and influenza VCR were analyzed across 21 regions in Italy. No correction for confounding was performed. Data up to May 2020 were used. | Moderate to strong, negative correlation between influenza vaccination and COVID-19 attributable mortality. ($r = -.5874$, $n = 21$, $P = .0051$) |
| Amato et al., 2020 | Ecological study | Influenza VCR and SARS-CoV-2 seroprevalence, hospitalization rates, ICU admissions as well as COVID-19 related mortality in the elderly (>65 years) in Italy were assessed. Correction for confounders was performed for several comorbidities, economic and environmental variables. COVID-19 related data were collected from March 2020 until June 2020. Influenza VCR was calculated based on extrapolation of the last five years' vaccination rates. | Negative correlation between influenza vaccination and SARS-CoV-2 seroprevalence, hospitalization rates, ICU admission as well as COVID-19 attributable mortality. ($R^2$ of 0.88, 0.82, 0.70 and 0.78 respectively) |
| Zanettini et al., 2020 | Ecological study | COVID-19 mortality in elderly (> 65 years) and influenza VCR were analyzed across all states of the USA. Correction for confounding was performed for several socio-economic-, demographic-, healthcare-, race- and comorbidity- related variables. Data from January 2020 until June 2020 were used. | Negative association between influenza vaccination and COVID-19 attributable mortality, where every 10% increase in influenza VCR, causes a 28% decrease in COVID-19 related death. (MRR = 0.72; 95%CI: 0.58-0.89) |
| Arokiaraj, 2020 | Ecological study | COVID-19 morbidity and mortality in elderly (> 65 years) and influenza VCR were analyzed across OECD countries. | Negative correlations between influenza vaccination and the COVID-19 epidemiological parameters. However, lacking of statistical analysis. |
| Lisewski et al., 2020 | Ecological study | COVID-19 incidence in elderly (> 65 years) and influenza VCR were analyzed across 29 OECD countries. | Positive correlation between influenza vaccination and COVID-19 attack rates. ($r = 0.58$; 95%CI: 0.27 to 0.78; $P = .001$) |
| EBMPHET consortium, 2020 | Ecological study | COVID-19 mortality in elderly (> 65 years) and influenza VCR were analyzed across Europe and the USA. Data up to May 2020 were used. No correction for confounding was performed. | Positive correlation between influenza vaccination and COVID-19 incidence in Europe ($r = 0.66 \pm 0.13$, $P = .000017$) and the USA ($r = 0.50 \pm 0.14$, $P < .05$). Positive correlation between influenza vaccination and COVID-19 attributable mortality in Europe ($r = 0.68 \pm 0.13$, $p = 0.000006$) and the USA ($r = 0.50 \pm 0.14$, $P < .05$). |
| Fink et al., 2020 | Cross-sectional study | Associations between COVID-19 related mortality, intensive care treatment, need of invasive respiratory support and influenza VCR were analyzed in 92.664 clinically and molecularly confirmed COVID-19 cases in Brazil. Correction for confounding was performed for several comorbidities, sociodemographic factors and healthcare facilities. | Positive correlation between influenza vaccination and COVID-19 attributed mortality (17% lower odds, 95%CI [0.75,0.89]). Positive correlation between influenza vaccination and need of intensive care treatment (8% lower odds, 95%CI [0.86,0.99]). Positive correlation between influenza vaccination and need of invasive respiratory support (18% lower odds, 95%CI [0.74,0.88]) |

ICU, intensive care unit; OECD, Organization for Economic Cooperation and Development; VCR, vaccination coverage rate.

Overall, the majority of the ecological data are leaning to a possible protective effect of the influenza vaccines. However, ecological studies also have several limitations. In this respect, there can be systematic differences in how countries and areas report disease, mortality and exposures. For example, in Italy any deceased person with a positive SARS-CoV-2 test is registered as a SARS-CoV-2 related death [28]. Besides this, information on confounding factors and effect modifiers can be missing and correction for confounders sometimes is impossible, causing over- or underestimation of outcomes. Wals et al. reviewed 18 studies in which influenza and its associations with other respiratory infections as well as COVID-19 were assessed. The conclusion was that live influenza vaccines are safe, however data on trivalent inactivated vaccines were not reassuring, followed by the suggestion to vaccinate with live vaccines when possible [29]. Aside from ecological studies, there is a recent cross sectional study conducted in Brazil, also supportive of a negative correlation between an inactivated trivalent influenza and COVID-19 attributed mortality (17% lower odds, 95%CI [0.75,0.89]), need of intensive care treatment (8% lower odds, 95%CI [0.86,0.99]) and need of invasive respiratory support (18% lower odds, 95%CI [0.74,0.88]). Correction for comorbidities, several sociodemographic factors and healthcare facilities was performed [30].

Many of these studies hypothesized that trained immunity may be the mechanism underlying these observations [21, 31-33]. The most extensively studied vaccine that induces trained immunity is BCG, which is currently being examined for its putative protective effects against COVID-19 duration and severity in several clinical trials (NCT04328441, NCT04348370, NCT04327206, NL8609). Although this property is usually assigned to live vaccines [34], whether influenza vaccination can also induce trained immunity was not known. In this study we also found that Vaxigrip Tetra® induces a trained immunity response towards both SARS-CoV-2 and the TLR4 ligand LPS, and in addition synergized with the trained immunity effects of BCG. The fast induction of cytokine responses at the beginning of the infection is crucial to decrease the viral load and prevent systemic inflammation. The amplified IL-6 response activates acute phase proteins, stimulates effector T-cell development along with antibody secretion, forming the linkage between innate and adaptive immunity, thus contributing to the clearance of the infection [35]. On the other hand, anti-inflammatory cytokines such as IL-1Ra are necessary to fine-tune the inflammation and counteract excessive inflammation. In our experimental setup we observed both, increased production of IL-6 paralleled with IL-1Ra, after stimulation with the influenza vaccine and BCG. This confirms that these cytokines might contribute to keeping a balance in the inflammatory status of the individual [36]. In addition, trained immunity is also known to be induced in NK cells, which play an important role in containing viral infections, among others through their production of IFN-γ. The increased IFN-γ produced after stimulation with the influenza vaccine and BCG, as shown here, can indicate the long-term functional reprogramming of NK cells, which then activate macrophages to further orchestrate the clearing of pathogens [37].

Our study also has important limitations. The database analysis performed in this study did not allow to correct for confounders, as we were not able to access data of individual influenza vaccination status in SARS-CoV-2 negative employees. An important confounder is the difference in the rate of the direct contact with patients between employees who developed SARS-CoV-2 or not, since this was the variable unevenly distributed among the two groups, with less direct patient contact in SARS-CoV-2 negative employees. However, earlier studies have reported that most of the SARS-CoV-2 infections in hospital personnel occur in society, rather than through patient contact in the hospitals [38-40]. Furthermore, we had no information on comorbidities in SARS-CoV-2 negative personnel or other exposures outside the hospital environment. Within the SARS-CoV-2 positive group, part of the individuals did not return the questionnaire which included information about comorbidities. These missing values were left out and could not be computed which can affect the analysis as well. Lastly, one cannot rule out healthy-vaccinee bias, and caution is always required when translating survey data into real-life conditions.

In conclusion, we provide observational data suggesting a potentially protective role of the quadrivalent inactivated influenza vaccine on COVID-19 incidence. In addition, we report first insights in the immunological mechanisms underlying these observations. We show that a quadrivalent inactivated influenza vaccine can induce trained immunity, and the plausible mechanisms through which an enhanced antiviral state is acquired after vaccination. Considering these data, and with at least several months more needed until a specific SARS-CoV-2 vaccine is available, influenza vaccination may contribute not only to reduction of influenza but also to the COVID-19-related burden on the healthcare system. While our data show that earlier influenza vaccination is safe in relation to a later SARS-CoV-2 infection, we recommend vaccination in the absence of active COVID-19, because of the theoretical possibility to induce a cytokine storm by an enhanced immune response if the vaccine is given during an active infection. Additionally, our data suggest that the presence of a previous BCG vaccination prior to the influenza vaccination could lead to enhanced responses and improved protection, raising the possibility to conduct clinical trials to asses this hypothesis.

## Data Availability

The data that support the findings of this study are available from the corresponding author, upon reasonable request.

## ACKNOWLEGDEMENTS

MGN was supported by a Spinoza Grant of the Netherlands Association for Scientific Research and an ERC Advanced Grant (no. 833247). PNO, LM and HS were supported by the Jürgen Manchot Foundation. We thank the Department of Occupational Health and Safety (Radboud University Medical Center, Nijmegen) for providing Vaxigrip Tetra®. We also thank Yuri Elsas (Radboud University, Nijmegen) for assistance in collecting the questionnaires.

## AUTHOR CONTRIBUTIONS

PAD and MGN designed the studies. PAD conducted the experiments, analysis and conceptualized the manuscript. All coauthors provided input on draft versions and approved the final version.

## COMPETING INTERESTS

The authors declare no competing interests.

## Notes

### Competing Interest Statement

The authors have declared no competing interest.

### Author Declarations

CMO regio Arnhem- Nijmegen

