## Supplementary figure 1 for "The effect of influenza vaccination on trained immunity: impact on COVID-19"

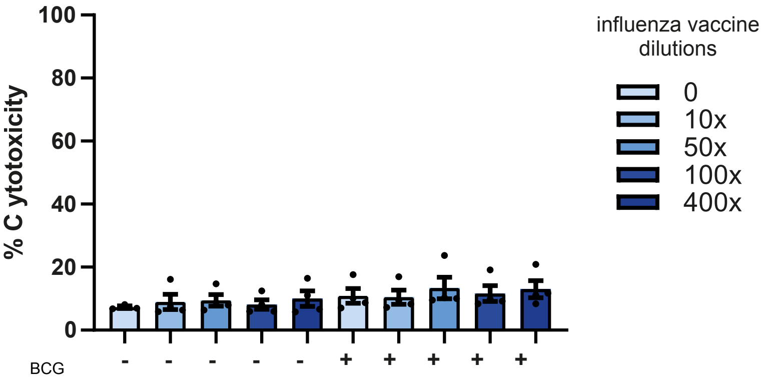


**Supplementary figure 1 – LDH assay**

PBMCs from healthy donors were trained with RPMI, 4 dilutions (10x, 50x, 100x, 400x) of the quadrivalent inactivated influenza vaccine (Vaxigrip Tetra®) alone or in presence of BCG. LDH concentrations were measured in fresh 24 h supernatants and calculated to percentages of cytotoxicity in relation to lysed cells. No significant differences were found among the various conditions (Wilcoxon matched pairs signed rank-test, $n$ = 3).
